# A pilot analysis of circulating cfRNA transcripts for the detection of lung cancer

**DOI:** 10.1101/2022.08.21.22279038

**Authors:** Chamindi Seneviratne, Amol C. Shetty, Xinyan Geng, Carrie McCracken, Jessica Cornell, Kristin Mullins, Feng Jiang, Sanford Stass

## Abstract

Lung cancers are the leading cause of cancer-related deaths worldwide. Studies have shown that non-small cell lung cancer (NSCLC) which constitutes majority of lung cancers, are significantly more responsive to early-stage interventions. However, the early stages are often asymptomatic, and current diagnostic methods are limited in their precision and safety. The cell-free RNAs (cfRNA) circulating in plasma (Liquid biopsies) offer non-invasive detection of spatial and temporal changes occurring in primary tumors since early stages. To address gaps in current cfRNA knowledgebase, we conducted a pilot study for comprehensive analysis of transcriptome-wide changes in plasma cfRNA in NSCLC patients. Total cfRNA was extracted from archived plasma collected from NSCLC patients (N=12), cancer-free former smokers (N=12) and non-smoking healthy volunteers (N=12). Plasma cfRNA expression levels were quantified by using a tagmentation-based library preparation and sequencing. The comparisons of cfRNA expression levels between patients and the two control groups revealed a total of 2357 differentially expressed cfRNA enriched in 123 pathways. Of these, 251 transcripts were previously reported in primary NSCLCs. A small subset of genes (N=5) was validated in an independent sample (N=50) using qRT-PCR. Our study provides a framework for developing blood-based assays for early detection of NSCLC and warrants further validation.

## 1. Introduction

Lung cancers are the leading cause of cancer-related deaths in both men and women in the U.S. and worldwide. Non-small cell lung cancer (NSCLC) constitutes approximately 84% of all lung cancer cases, and consists of two main histological subtypes: adenocarcinoma (AC) and squamous cell carcinoma (SCC) [1]. The main risk factor for developing NSCLC is smoking, which is preventable, yet highly prevalent with over a billion smokers around the world [2]. Moreover, smoking, and other environmental pollutants interact with biological factors such as aging and genetic risk variants to increase disease burden [3-6]. Furthermore, the NSCLC risk has been shown to correlate positively with severity and duration of smoking, and negatively with time since smoking cessation [7, 8].

Because the lung cancers are often asymptomatic in early stages, most patients are diagnosed at advanced stages resulting in only about 15-20% of patients surviving 5 years after the diagnosis [6]. The early-stage NSCLC are more responsive to treatment [9], and therefore, crucial to reduce mortality. At present, the only recommended diagnostic method for NSCLC is detection of pulmonary nodules (PNs) with low dose computed tomography (LDCT) [10]. In fact, based on data from Cancer Intervention and Surveillance Modeling Network (CISNET), the US Preventive Services Task Force (USPSTF) recommended annual screening of adults aged 50 to 80 years of age with a smoking history of 20 or more pack-years, who currently smoke or quit smoking within the past 15 years [11]. This 2021 USPSTF recommendation (A-50-80-20-15) was updated to expand the population eligible for LDCT screening over the previous 2013 USPSTF recommendation that required smoking history of 30 or more pack-years (A-50-80-30-15). The LDCT has high negative predictive values, moderate sensitivity and specificity, and low positive predictive values [12]. A recent meta-analysis corresponding to data from 84,558 participants who had a smoking history 15 or more pack-years indicated a 17% relative reduction in mortality in the group screened with LDCT compared to the control group [12]. Despite these encouraging statistics, there are several important limitations to using LDCT for NSCLC diagnosis. For example, the high false-positive rates can lead to further testing of benign PNs with invasive diagnostic and therapeutic procedures such as serial CTs, biopsy and surgery that carry their own morbidity. The invasive procedures are reported to be performed in 44% of smokers with indeterminate PNs that have roughly 5% probability of malignancy, and 35% of surgical resections are ultimately determined to be benign diseases [13]. Another concern is the exposure to radiation with repeated LDCT. Statistical modelling has predicted 1 death for every 13.0 lung-cancer related deaths avoided by LDCT with 2021 USPSTF recommendations, which was a 2% worsening compared to risk associated with 2013 USPSTF recommendations[11]. Considering these factors, it is clinically important to develop noninvasive biomarkers to distinguish malignant from benign PNs facilitating positive screening results on LDCT.

Recently, the concept of liquid biopsies has garnered excitement among the scientific community for its’ potential to provide real-time information on spatial and temporal changes in tumor markers in an easily obtained peripheral blood sample [14]. Several types of biomarkers have been explored in liquid biopsies as potential diagnostics with mixed results. Circulating tumor DNAs (ctDNA) have over 90% sensitivity and specificity for NSCLC diagnosis in patients with stage II–IV NSCLC, but around 50% in patients with stage I NSCLC when shedding rates are low [15]. Analysis of mutations in ctDNA has also been reported to have lower sensitivity and specificity in early-stage NSCLC [16]. Therefore, analysis of ctDNA-mutations or quantities appear to be more suitable for therapeutic and disease monitoring in NSCLC patients, rather than the early detection. In contrast, tumors with low shedding rates add cell-free RNAs (cfRNA) to blood circulation presenting with the opportunity to identify the over-expressed, tumor-specific, and tumor-derived RNA signals in the blood [17] at early stages, potentially facilitating high rates of patients able to receive curative surgical resections. Studies have also shown that cfRNA could complement ctDNA and thus improve early diagnosis [18]. The studies of cfRNA have mainly focused on either microRNAs (miRNA) or a small number of known cancer-related messenger RNAs (mRNA)[19-21]. Moreover, the published studies have used large amounts of plasma – up to 4-5ml, for cfRNA extraction for expression analyses limiting its potential clinical use. We have conducted a pilot study to explore the ability to detect cfRNA signatures of NSCLC, particularly of the genes that were previously reported to be differentially expressed in lung cancer primary tissue biopsies, compared to both cancer-free smokers and healthy non-smokers.

## 2. Materials and Methods

### Study design

In this pilot study, we first compared the expression levels of plasma cfRNA obtained from SCC and AC patients (N=12; cases) and cancer-free former smokers (N=12; control_smokers). As all patients in the case group were also heavy smokers, we included a second control group of non-smoking healthy individuals (N=12; control_healthy) to exclude differentially expressed cfRNAs associated with smoking, rather than pathological processes underlying NSCLC. Each participant provided whole blood samples as part of an umbrella protocol approved by the Institutional Review Board of the University of Maryland Baltimore [UMB IRB protocol ID: HP-00040666] and the Veterans Affairs Maryland Health Care System. All participants provided written informed consent to participate in research conducted at the University of Maryland Medical Center and the Baltimore VA Medical Center. Diagnosis of lung cancer was established by pathologic examination of tissues obtained via surgery or biopsy. Histological diagnosis was made on bronchoscopic biopsy specimens and thoracotomy according to the World Health Organization (WHO) categories. The NSCLC stage classification was based on WHO classification and the International Association for the Study of Lung Cancer staging system. The smokers consisted of former smokers who had a minimum smoking history of 30-pack years and quit within the past 15 years. Exclusion criteria were similar to Leng et al. 2017 [8]. Demographic and clinical characteristics of the cohorts are presented in Table 1.

**Table 1:** Demographic and clinical characteristics

|  | Cases | Control_<br>Smokers | Control_<br>Healthy | p-value<br>Cases vs. |  | p-value<br>Smokers vs.<br>Healthy |
| --- | --- | --- | --- | --- | --- | --- |
|  |  |  |  | Control_<br>Smokers | Control_<br>Healthy |  |
| Discovery cohort (N=36): |  |  |  |  |  |  |
| Sample size | 12 | 12 | 12 |  |  |  |
| Age (mean, (SD)) | 67.17 (8.99) | 68.44 (10.01) | 40.17 (4.99) | 0.728 | <0.0001 | <0.0001 |
| Gender<br>(Male, N (%)) | 11 (91.67) | 9 (75) | 7 (58.33) | 0.3144 | 0.0480 | 0.3144 |
| Race<br>(Caucasian, N (%)) | 5 (4.67) | 5 (4.67) | 5 (4.67) | ns | ns | ns |
| Stage |  |  |  |  |  |  |
| Stage I (N) | 7 (AC=5) |  |  |  |  |  |
| Stage II (N) | 4 (AC=1) |  |  |  |  |  |
| Stage III-IV (N) | 1 (AC=0) |  |  |  |  |  |
| Histological type |  |  |  |  |  |  |
| AC (N) | 6 |  |  |  |  |  |
| SCC (N) | 6 |  |  |  |  |  |
| Average plasma<br>volumes used (ml) | 1.6 | 1.6 | 1.54 | ns | ns | ns |
| Validation cohort (N=50): |  |  |  |  |  |  |
| Sample size | 25 | 18 | 7 |  |  |  |
| Age (mean, (SD)) | 64.60 (8.97) | 61.28 (10.23) | 58.14(17.38) | ns | ns | ns |
| Gender<br>(Male, N (%)) | 19 (76.00) | 13 (72.22) | 5 (71.43) | ns | ns | ns |
| Race<br>(Caucasian, N (%)) | 52 (50.99) | 11 (61.11) | 3 (60.00) | ns | ns | ns |
| Stage |  |  |  |  |  |  |
| Stage I (N) | 4 (AC=2) |  |  |  |  |  |
| Stage II (N) | 2 (AC=0) |  |  |  |  |  |
| Stage III-IV (N) | 9 (AC=8) |  |  |  |  |  |
| Missing data | 10 (AC=7) |  |  |  |  |  |
| Histological type |  |  |  |  |  |  |
| AC (N) | 13 |  |  |  |  |  |
| SCC (N) | 12 |  |  |  |  |  |
| Average plasma<br>volumes used (ml) | 0.5 | 0.5 | 0.5 |  |  |  |

ns – not significant ( $p>0.05$ ); AC – adenocarcinoma; SCC – squamous cell carcinoma.

### Sample preparation and sequencing

Archived plasma samples (volumes given in Table 1) prepared from 3-6 mL of whole blood collected into tubes containing EDTA were thawed at 37°C and centrifuged at 16,000 x G for 30 minutes at 4°C to remove any cellular components in plasma. The supernatant was extracted and centrifuged again at 13,000 x G for 30 minutes at 4°C, and store at −80°C until day of cfRNA extractions. Quality control procedures for plasma sample preparation were similar to our earlier study[22]. The cfRNA was extracted from archived plasma using the miRNeasy® Serum/Plasma Advanced Kit (Qiagen) according to manufacturer’s guidelines and were tested for RNA integrity using an *Agilent* bioanalyzer system. Libraries were prepared using a tagmentation-based method consisting of a two-step probe-assisted exome enrichment for cfRNA detection (Illumina, Inc, San Diego, CA) [23]. An Illumina Exome enrichment panel that included > 425,000 probes (oligos), each constructed against the NCBI37/hg19 reference genome, covering > 98% of the RefSeq exome was used to pool libraries with target cfRNA of interest. The probe set was designed to capture > 214,000 targets, spanning 21,415 genes of interest. Probes hybridized to target libraries were captured according to protocol and amplified using a 19-cycle PCR program. Enriched libraries were then purified with magnetic beads and were then sequenced using a NovaSeq 6000 system (Illumina, Inc), at a sequencing depth of 100 million reads at 100bp PE length sequences.

### Sequencing data analyses

Raw sequence reads generated for each sample were analyzed using the CAVERN analysis pipeline [24]. Read quality was assessed using the FastQC toolkit to ensure good quality reads for downstream analyses. Reads were aligned to the Human reference genome GRCh38 (available from *Ensembl* repository) using HISAT2, a fast splice-aware aligner for mapping next-generation sequencing reads [25]. Reads were aligned using default parameters to generate the alignment BAM files. Read alignments were assessed to compute gene expression counts for each gene using the HTSeq count tool [26] and the Human reference annotation (GRCh38). The raw read counts were normalized for library size and dispersion of gene expression. The normalized counts were utilized to assess differential cfRNA expression between conditions using DESeq2. P-values were generated using Wald test implemented in DESeq2 and then corrected for multiple hypothesis testing using the Benjamini-Hochberg correction method [27]. Significant differentially expressed cfRNA between conditions were determined using a false discovery rate (FDR) of 5% and a minimum absolute log2(fold-change) of 1.

### Quantitative RT-PCR (qRT-PCR) for validation of a subset of cfRNA

Based on findings from sequencing data analysis, we selected five differentially expressed protein coding genes as listed in Table 1 and detailed below in the results section for validation assays. We assessed the abundance of cfRNA for the five selected genes using qRT-PCR in an independent set of plasma samples from 25 cases (AC=13; SCC=12) and 25 controls (control_smokers=18; control_healthy=7). The demographic and clinical characteristics of the validation cohort are presented in Table 1. Total cfRNA was extracted from archived plasma (500ul per sample) using the same protocol described above for the discovery cohort. A mixture of three commercially available RNA spike-ins (miRNAs UniSp2, UniSp4 and UniSp5) from the *RNA Spike-In Kit, For RT* were added to plasma samples according to the manufacture’s protocol (Qiagen, Germantown, MD, USA) prior to extraction of cfRNA to control for cfRNA isolation across samples. The extracted total cfRNA samples were then split into equal volumes for cDNA synthesis and subsequent mRNA quantification and detection of the three-miRNA spike-ins in parallel. We used *miRCURY LNA RT* and *miRCURY LNA SYBR Green PCR* kits (Qiagen) for reverse transcription and qPCR of spike-in miRNAs, and the *QuantiTect® Reverse Transcription* and *QuantiTect SYBR Green RT-PCR* kits (Qiagen) for reverse transcription and qPCR of the selected protein coding genes. All qPCR reactions were performed in triplicates with 1:10 cDNA dilutions on a Bio-Rad CFX real-time PCR detection system (Bio-Rad, Hercules, California, USA), according to protocols associated with each kit. As stable endogenous reference genes for quantifying circulating mRNA in plasma samples have not been established in literature and normalizing to a global mean of all expressed mRNA was not applicable to the analysis of five genes, we opted for not using a reference gene in this pilot study. We also explored the possibility of using GAPDH - the commonly used endogenous reference gene for cellular mRNA and did not detect any amplification. Therefore, we adopted a method of first assessing the between sample variability using three spike-ins to identify outlier samples, and then perform qRT-PCR for the five selected genes excluding outliers. Two tailed t-tests in GraphPad Prism software (San Diego, CA, USA) were performed for statistical comparisons.

## 3. Results

### cfRNA processing and quality control

cfRNA was extracted from all 36 samples at mean concentrations of 0.111 ng/ul in cases, 0.085 ng/ul in control_smokers, and 0.151ng/ul in control_healthy group. The RNA integrity numbers (RIN) ranged from 1 to 5.3. All samples had sequence reads that mapped >80% to the reference sequence and mapped to exonic regions. Total Gene Abundance ranged from approximately 10 to 70 million. Of these genes, 0.5-10% were Hb coding genes, 0.5-20% mitochondrial genes, <0.03% ribosomal RNA (rRNA) genes, and up to 4% were other non-coding RNA (ncRNA) genes. Amongst protein coding genes, the most abundant were actin, myosin, platelet-specific genes, and pseudogenes.

### Identification of differentially expressed cfRNA between cases and controls

Differential expression of cfRNA was analyzed after excluding Hb, Mitochondrial and rRNA transcripts. As shown in Figure 1.1, a total of 1,905 (X+Y+Z) cfRNA were identified to be differentially expressed in plasma samples from cases compared to the two control groups. Of these, 2 cfRNA (LINC01956 and TAS2R16) were differentially expressed in opposite directions in cases compared to control_smokers and control_healthy groups, and therefore we have included theses in both X and Z categories in Figure 1.1: Both cfRNA were downregulated compared to control_smokers and upregulated compared to control_healthy group. Another 1,377 (B+C+D in Figure 1.1) cfRNA that were detected in cases, were differentially expressed in the same direction in cancer-free smokers. The volcano plots for comparison of cfRNA differential expression between cases and controls are presented in Figures 2.1 and 2.2.

**Figure 1:**
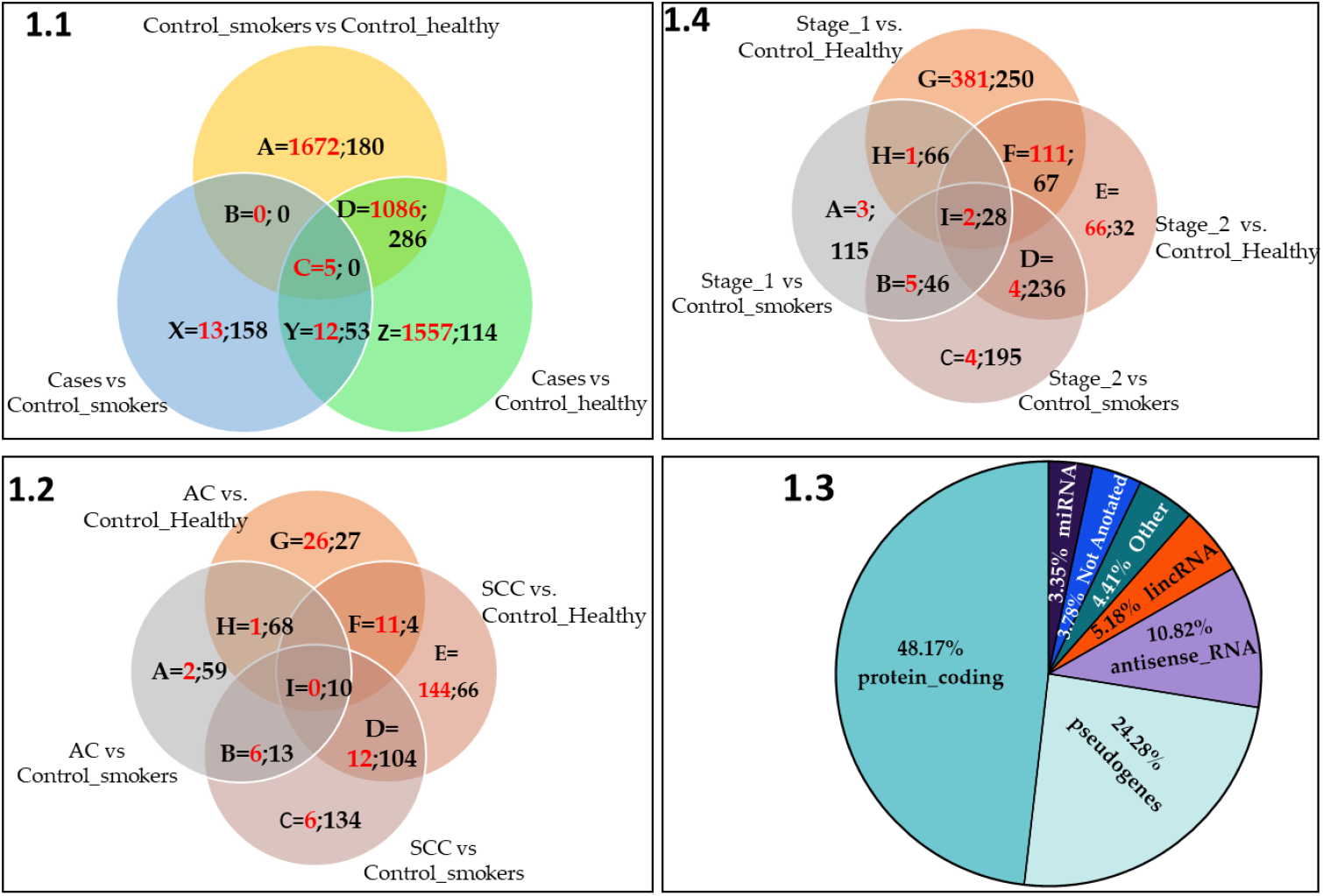
Distribution of NSCLC associated cfRNA. **1.1**: cfRNA in plasma samples from cases **1.2**: cfRNA in subtypes AC and SCC. **1.3**: Distribution of NSCLC associated cfRNA within functional categories. The most common pseudogene subcategories were, processed_pseudogenes (17.99%), unprocessed_pseudogenes (2.89%), transcribed_unprocessed_pseudogenes (1.82%) and other subtypes were present <1%. The “Other” category included the following subcategories at less than 1% abundance: IG_V_genes, snoRNA, processed_transcripts, TR_V_genes, TR_J_genes, sense_intronic, misc_RNA, scaRNA, sense_overlapping, IG_C_genes, TR_C_genes, 3prime_overlapping_ncRNA, IG_J_genes, TEC, and TR_D_genes. **1.4:** cfRNA within categories based on NSCLC stage. The numbers presented in red and black color font in Figures 1.1, 1.2 and 1.3 represent up- and down-regulated genes, respectively.

**Figure 2:**
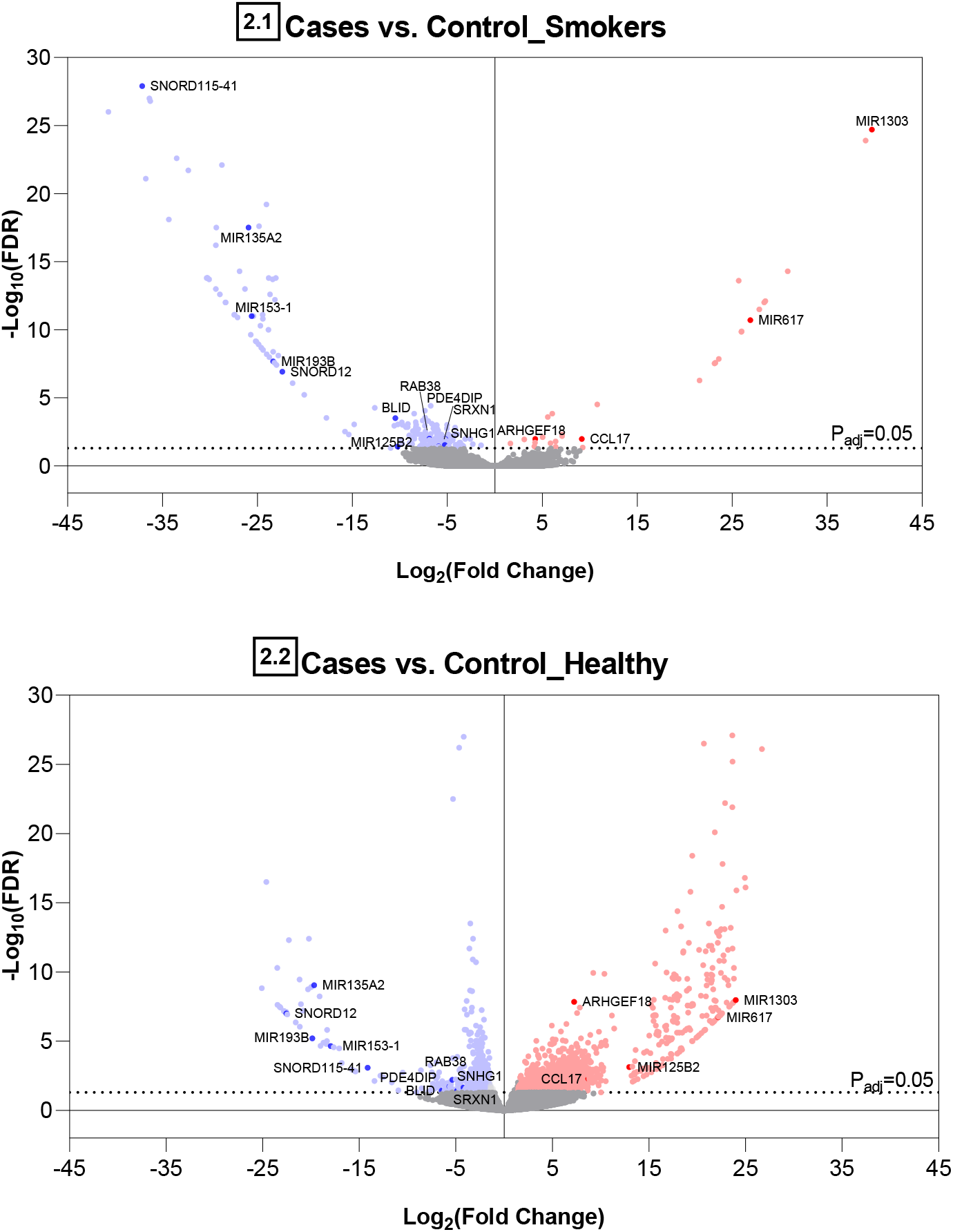
Volcano plots for (1) cases vs. smokers with benign PN (Figure 2.1), and (2) cases vs. healthy non-smokers (Figure 2.2). The horizontal dotted lines indicate an adjusted p-value of 0.05. The dots are colored blue or red if classified as down-or upregulated, respectively, using a threshold of log 2-fold change of −1 and 1.

#### Statistical power analysis

post-hoc power analysis revealed that the sample of 12 cases and 24 controls afforded a 78.5% power to detect differentially expressed genes with 2-fold effect size using a 5% false discovery rate.

### Exploratory subgroup analysis

We performed two subgroup analyses exploring differentially expressed cfRNA between (1) subtypes of cases - AC vs. SCC, and (2) based on NSCLC stage – stages 1 vs. 2, compared to both control groups irrespective of their statistical significance in the combined case group. Figure 1.2 presents all cfRNA within each subtype category excluding DEGs shared with cancerfree smokers (i.e., comparisons between control_smokers and control_healthy groups). Of these, a total of 452 cfRNA (64.3% all DEGs in Figure 1.2) were not detected in the combined cases (X+Y+Z in Figure 1.1), but uniquely differentially expressed in either AC or SCC, or both but in differing directions. As depicted in Figure 1.3, nearly half of all 2357 total cfRNA (1905+452) were functional protein coding genes (Figure 1.3). All cfRNA included in Figure 1 are listed in Supplementary Table 1. Similarly, Figure 1.4 presents cfRNA comparisons between NSCLC stages I and II, excluding cfRNA shared with cancer-free smokers. Comparisons with other NSCLC stages were not possible as we had only one sample from a patient diagnosed with stage III and none for stage IV. Results indicated that 1,075 genes to be expressed in plasma from patients who had stage 1 NSCLC (A+B+H+I+G+F in Figure 1.4), out of which 259 were common to both stages I and II. As both subgroup analyses had small numbers of patients within each category (Table 1), these findings should only be considered as exploratory.

**Figure 3:**
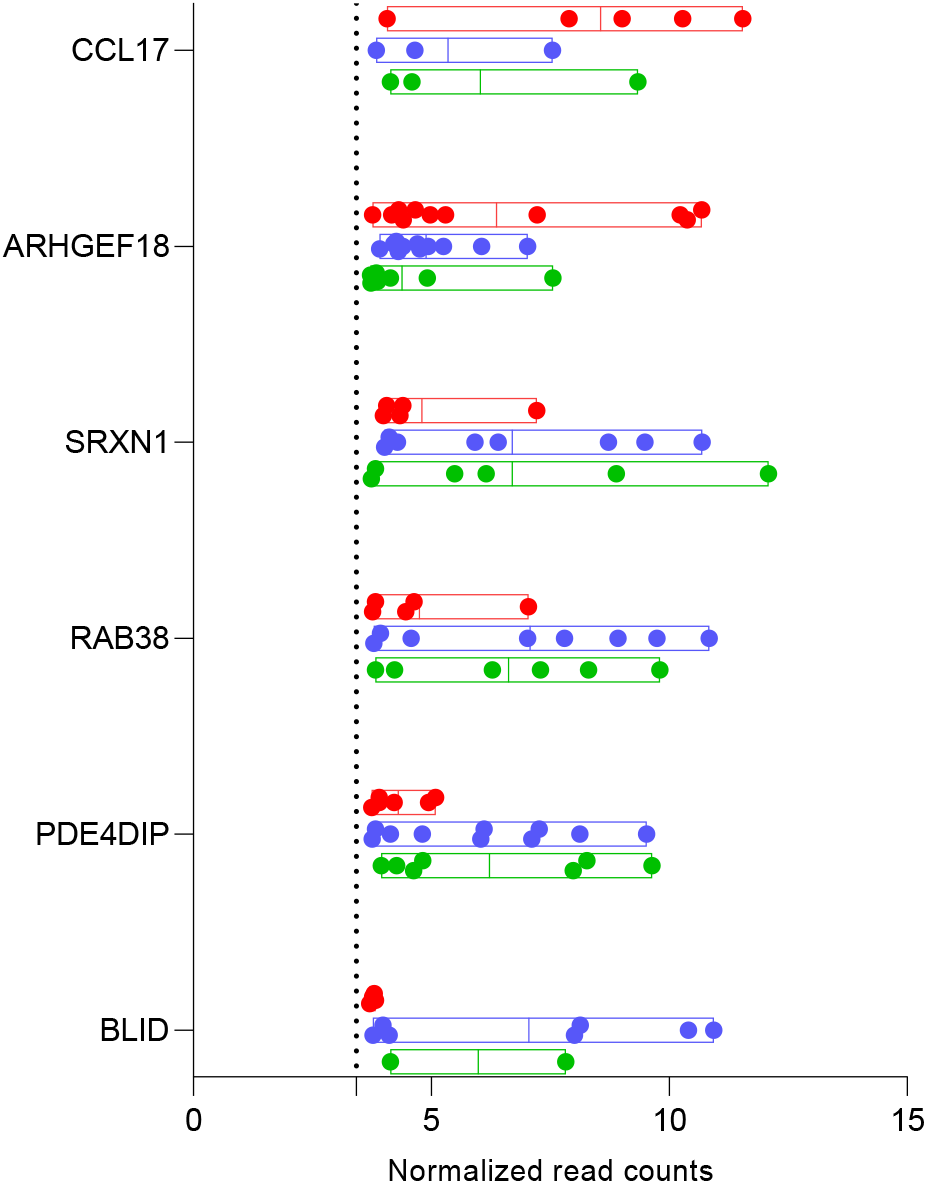
Distribution of read counts across individual samples for cfRNA of replicated protein coding genes. Each dot represents cfRNA read counts for a given gene within individual samples. Red –genes in cases; blue – smokers with benign PN; green – healthy non-smokers. The dotted line represents the threshold for detecting read counts which was set at 3.4298.

**Figure 4:**
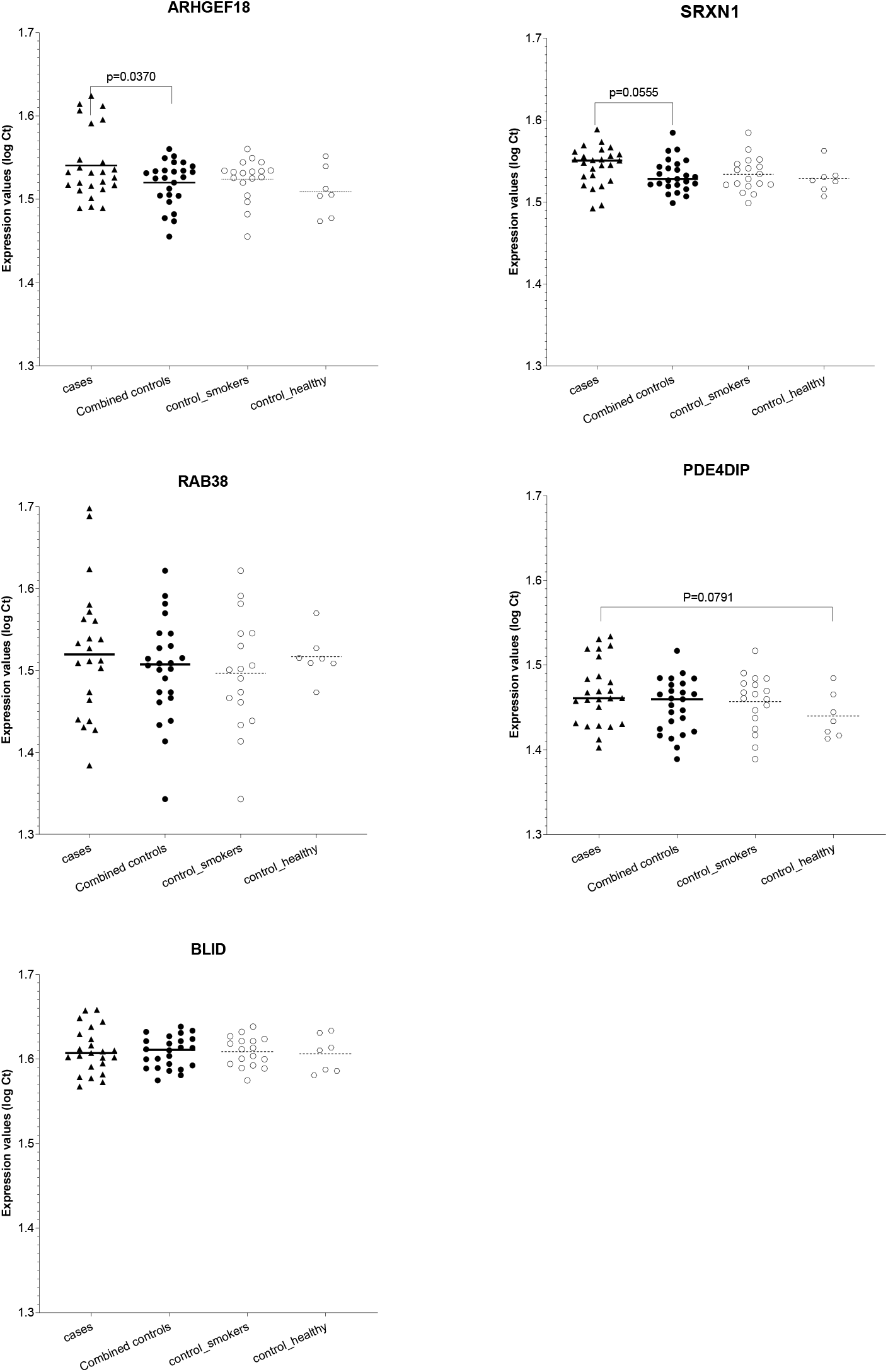
qRT-PCR analysis of changes in the expression levels of cfRNA of selective protein coding genes in an independent sample. Each symbol represents log transformed Ct values of mRNA levels within each sample averaged across three technical repeats. The horizontal lines represent mean expression levels within each group.

### Literature review to identify DEGs previously reported in primary NSCLC biopsies

We performed an exhaustive review of all published studies listed on National Center for Biotechnology Information (NCBI)’s database for gene-specific information, using gene IDs for each of the 2357 identified DEGs. Studies reporting DEGs in primary NSCLC biopsies were identified and are referenced in *Supplementary Table 1*. Our literature review showed that 10.65% of total DEGs (N=251 of 2357) have been reported in primary tumor biopsies from NSCLC patients in published studies. Majority of these replicated genes were mRNA transcripts of protein coding genes (N=174; 69.32%), while some (N=45; 17.92%) were miRNA. Next, to assess inter-patient variation in cfRNA transcript abundance within each group (i.e., combined cases, control_smokers and control_healthy), we evaluated whether the transcripts were expressed above detectable levels, and then calculated coefficients of variation (%CV) within a group for each gene. Of the total 174 replicated protein coding genes identified in this study, 78.97% were expressed above threshold in cases and 88% had <50% CV for each replicated gene (Supplementary Table 1). Fifteen cfRNA that were differentially expressed in cases compared to both control groups (category “Y” in Figure 1.1) and reported in primary NSCLC tissue biopsies are listed in Table 2. The distribution of these 15 replicated cfRNA that were differentially expressed in cases compared to the two control groups are marked in volcano plots presented in Figure 2.1 and Figure 2.2. Of the six replicated protein coding genes, all but CCL17 were expressed with <50% CV in samples within cases (Table 2 and Figure 3). Therefore, we selected the five genes (i.e., ARHGEF18, SRXN1, RAB38, PDE4DIP, and BLID) for further validation in an independent cohort.

**Table 2:**
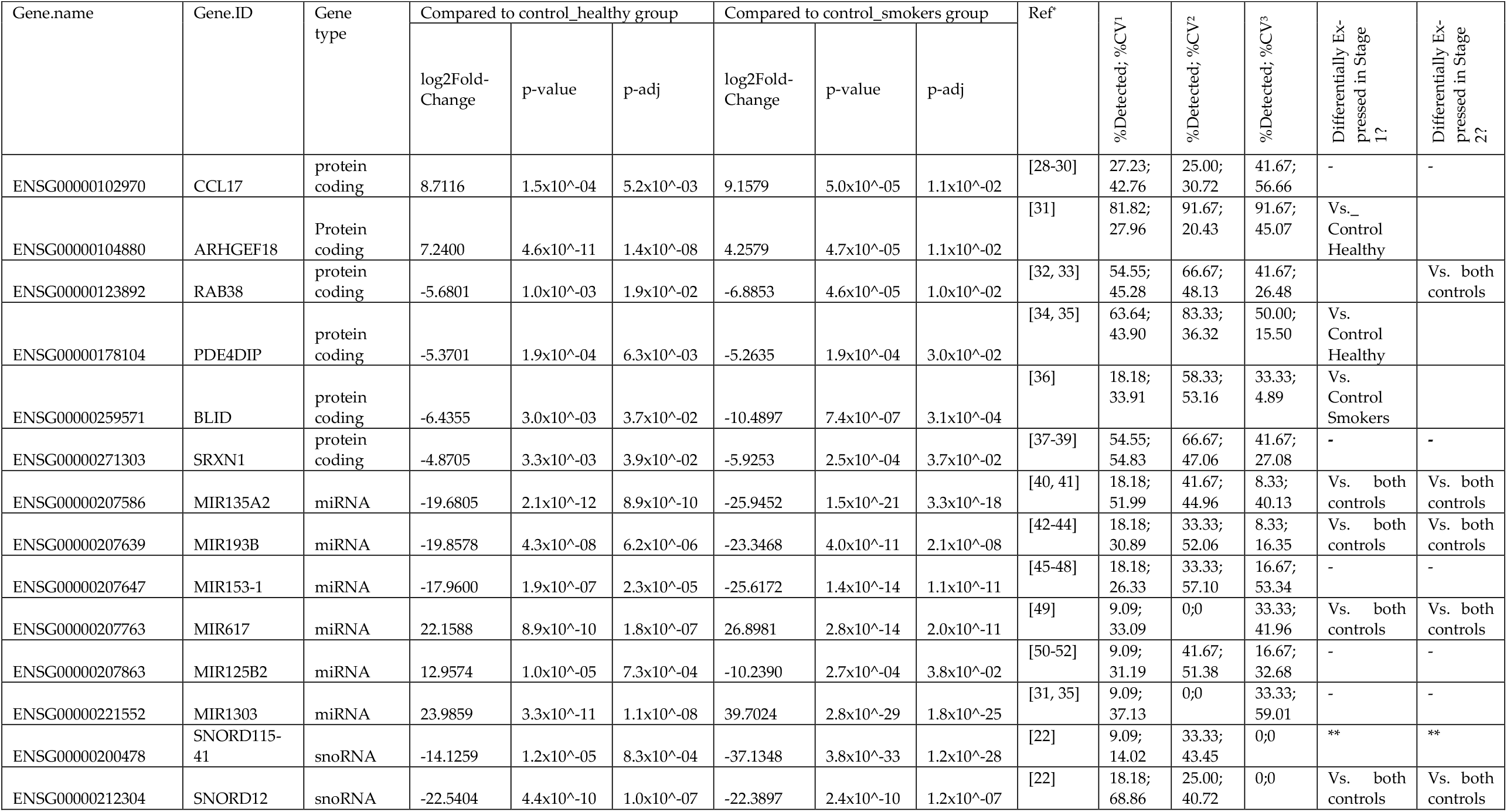
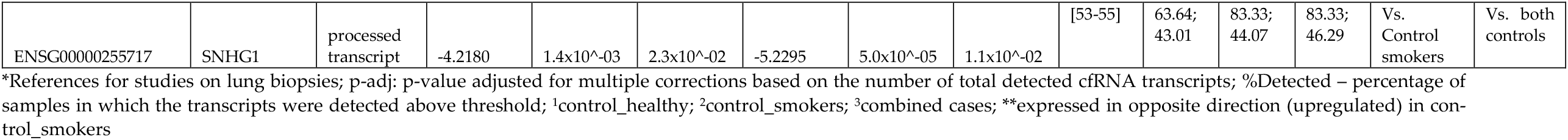
cfRNA differentially expressed in cases compared to both control groups and confirmed by published studies

| Gene.name | Gene.ID | Gene type | Compared to control_healthy group |  |  | Compared to control_smokers group |  |  | Ref<sup>a</sup> | %Detected; %CV<sup>1</sup> | %Detected; %CV<sup>2</sup> | %Detected; %CV<sup>3</sup> | Differentially Expressed in Stage 1? | Differentially Expressed in Stage 2? |
| --- | --- | --- | --- | --- | --- | --- | --- | --- | --- | --- | --- | --- | --- | --- |
|  |  |  | log2Fold-Change | p-value | p-adj | log2Fold-Change | p-value | p-adj |  |  |  |  |  |  |
| ENSG00000102970 | CCL17 | protein coding | 8.7116 | 1.5x10<sup>-04</sup> | 5.2x10<sup>-03</sup> | 9.1579 | 5.0x10<sup>-05</sup> | 1.1x10<sup>-02</sup> | [28-30] | 27.23; 42.76 | 25.00; 30.72 | 41.67; 56.66 | - | - |
| ENSG00000104880 | ARHGEF18 | Protein coding | 7.2400 | 4.6x10<sup>-11</sup> | 1.4x10<sup>-08</sup> | 4.2579 | 4.7x10<sup>-05</sup> | 1.1x10<sup>-02</sup> | [31] | 81.82; 27.96 | 91.67; 20.43 | 91.67; 45.07 | Vs. Control Healthy |  |
| ENSG00000123892 | RAB38 | protein coding | -5.6801 | 1.0x10<sup>-03</sup> | 1.9x10<sup>-02</sup> | -6.8853 | 4.6x10<sup>-05</sup> | 1.0x10<sup>-02</sup> | [32, 33] | 54.55; 45.28 | 66.67; 48.13 | 41.67; 26.48 |  | Vs. both controls |
| ENSG00000178104 | PDE4DIP | protein coding | -5.3701 | 1.9x10<sup>-04</sup> | 6.3x10<sup>-03</sup> | -5.2635 | 1.9x10<sup>-04</sup> | 3.0x10<sup>-02</sup> | [34, 35] | 63.64; 43.90 | 83.33; 36.32 | 50.00; 15.50 | Vs. Control Healthy |  |
| ENSG00000259571 | BLID | protein coding | -6.4355 | 3.0x10<sup>-03</sup> | 3.7x10<sup>-02</sup> | -10.4897 | 7.4x10<sup>-07</sup> | 3.1x10<sup>-04</sup> | [36] | 18.18; 33.91 | 58.33; 53.16 | 33.33; 4.89 | Vs. Control Smokers |  |
| ENSG00000271303 | SRXN1 | protein coding | -4.8705 | 3.3x10<sup>-03</sup> | 3.9x10<sup>-02</sup> | -5.9253 | 2.5x10<sup>-04</sup> | 3.7x10<sup>-02</sup> | [37-39] | 54.55; 54.83 | 66.67; 47.06 | 41.67; 27.08 | - | - |
| ENSG00000207586 | MIR135A2 | miRNA | -19.6805 | 2.1x10<sup>-12</sup> | 8.9x10<sup>-10</sup> | -25.9452 | 1.5x10<sup>-21</sup> | 3.3x10<sup>-18</sup> | [40, 41] | 18.18; 51.99 | 41.67; 44.96 | 8.33; 40.13 | Vs. both controls | Vs. both controls |
| ENSG00000207639 | MIR193B | miRNA | -19.8578 | 4.3x10<sup>-08</sup> | 6.2x10<sup>-06</sup> | -23.3468 | 4.0x10<sup>-11</sup> | 2.1x10<sup>-08</sup> | [42-44] | 18.18; 30.89 | 33.33; 52.06 | 8.33; 16.35 | Vs. both controls | Vs. both controls |
| ENSG00000207647 | MIR153-1 | miRNA | -17.9600 | 1.9x10<sup>-07</sup> | 2.3x10<sup>-05</sup> | -25.6172 | 1.4x10<sup>-14</sup> | 1.1x10<sup>-11</sup> | [45-48] | 18.18; 26.33 | 33.33; 57.10 | 16.67; 53.34 | - | - |
| ENSG00000207763 | MIR617 | miRNA | 22.1588 | 8.9x10<sup>-10</sup> | 1.8x10<sup>-07</sup> | 26.8981 | 2.8x10<sup>-14</sup> | 2.0x10<sup>-11</sup> | [49] | 9.09; 33.09 | 0;0 | 33.33; 41.96 | Vs. both controls | Vs. both controls |
| ENSG00000207863 | MIR125B2 | miRNA | 12.9574 | 1.0x10<sup>-05</sup> | 7.3x10<sup>-04</sup> | -10.2390 | 2.7x10<sup>-04</sup> | 3.8x10<sup>-02</sup> | [50-52] | 9.09; 31.19 | 41.67; 51.38 | 16.67; 32.68 | - | - |
| ENSG00000221552 | MIR1303 | miRNA | 23.9859 | 3.3x10<sup>-11</sup> | 1.1x10<sup>-08</sup> | 39.7024 | 2.8x10<sup>-29</sup> | 1.8x10<sup>-25</sup> | [31, 35] | 9.09; 37.13 | 0;0 | 33.33; 59.01 | - | - |
| ENSG00000200478 | SNORD115-41 | snoRNA | -14.1259 | 1.2x10<sup>-05</sup> | 8.3x10<sup>-04</sup> | -37.1348 | 3.8x10<sup>-33</sup> | 1.2x10<sup>-28</sup> | [22] | 9.09; 14.02 | 33.33; 43.45 | 0;0 | ** | ** |
| ENSG00000212304 | SNORD12 | snoRNA | -22.5404 | 4.4x10<sup>-10</sup> | 1.0x10<sup>-07</sup> | -22.3897 | 2.4x10<sup>-10</sup> | 1.2x10<sup>-07</sup> | [22] | 18.18; 68.86 | 25.00; 40.72 | 0;0 | Vs. both controls | Vs. both controls |
| ENSG00000255717 | SNHG1 | processed transcript | -4.2180 | 1.4x10<sup>-03</sup> | 2.3x10<sup>-02</sup> | -5.2295 | 5.0x10<sup>-05</sup> | 1.1x10<sup>-02</sup> | [53-55] | 63.64; 43.01 | 83.33; 44.07 | 83.33; 46.29 | Vs. Control smokers | Vs. both controls |

\*References for studies on lung biopsies; p-adj: p-value adjusted for multiple corrections based on the number of total detected cfRNA transcripts; %Detected – percentage of samples in which the transcripts were detected above threshold; <sup>1</sup>control\_healthy; <sup>2</sup>control\_smokers; <sup>3</sup>combined cases; \*\*expressed in opposite direction (upregulated) in control\_smokers

### Quantitative RT-PCR (qRT-PCR) for validation of replicated cfRNA of protein coding genes

While all listed genes in Table 2 are reported to underly pathophysiology of NSCLC, we specifically selected the protein coding genes for our initial validation as the circulating mRNA were the most abundant type of cfRNA present in our discovery cohort, and cf-mRNA are relatively less characterized in literature despite their biological relevance. Expression data for the three spike-ins in all 50 samples are presented in Supplementary Figure 1. As UniSp2, UniSp4 and UniSp5 were detected in all samples, we assessed cf-mRNA for the five genes in all 50 samples without excluding any. As shown in Figure 4, our findings indicated that three of the five tested genes differentially expressed between cases and controls. ARHGEF18 showed a nominally significant downregulation (i.e., higher Ct values) in cases (p=0.037), and SRXN1 showed a trend towards downregulation in cases (p=0.056) compared to the combined control group. PDE4DIP showed a trend towards downregulation in cases compared only to the healthy non-smokers (p=0.079). The other two genes-RAB38 and BLID, did not show statistically significantly expressed cfRNA levels between cases and controls.

### Gene ontology (GO) enrichment analysis of differentially expressed cfRNA

The unbiased pathway analysis with cfRNA for the differentially expressed genes included in each category of Figure 1.1 revealed 123 significantly enriched pathways across the three comparison groups. The cases compared to control_smokers had one significantly enriched pathway that was also detected in cancer-free smokers; GO:0010629 (negative regulation of gene expression) with 286 cfRNA in control_smokers vs control_healthy group (adjusted p= 0.0041) and 24 cfRNA in cases vs control_smokers group (adjusted p= 5.98E-05). However, at an individual gene level, only 2 cfRNA (MIR874 and MIR551B) in GO:0010629 were common to the 2 groups, both in terms of direction and type. The cases vs control_smokers and cases vs control_healthy comparisons did not share any significantly enriched pathways. Eighty-five pathways were commonly enriched in cases and cancer-free smokers when each group was compared with the control_healthy. Details of the 37 pathways that were uniquely enriched in cases compared with both control groups include general mechanisms underlying cancer biology and are presented in Table 3 below. The gene IDs for cfRNA enriched within these pathways are listed in Supplementary Table 2.

**Table 3:** Enriched pathways in cases compared to the two control groups

| ID | Description of pathway | Gene ratio | p-value | p-adjust |
| --- | --- | --- | --- | --- |
| **Cases vs. control_healthy:** |  |  |  |  |
| GO:0001501 | skeletal system development | 71/1685 | 8.5591E-05 | 0.016903 |
| GO:0005125 | cytokine activity | 42/1656 | 6.6270E-05 | 0.022704 |
| GO:0005198 | structural molecule activity | 105/1656 | 2.0920E-05 | 0.009907 |
| GO:0007186 | G protein-coupled receptor signaling | 179/1685 | 5.2099E-06 | 0.002827 |
| GO:0007200 | phospholipase C-activating G protein-coupled receptor signaling | 22/1685 | 4.5649E-05 | 0.011269 |
| GO:0007399 | nervous system development | 258/1685 | 0.0003 | 0.037022 |
| GO:0008154 | actin polymerization or depolymerization | 34/1685 | 0.0004 | 0.046481 |
| GO:0009888 | tissue development | 248/1685 | 1.0524E-06 | 0.001336 |
| GO:0009953 | dorsal/ventral pattern formation | 19/1685 | 0.0003 | 0.044695 |
| GO:0010454 | negative regulation of cell fate commitment | 7/1685 | 2.6616E-05 | 0.007885 |
| GO:0019958 | C-X-C chemokine binding | 5/1656 | 2.3133E-05 | 0.009907 |
| GO:0030545 | receptor regulator activity | 85/1656 | 2.1418E-06 | 0.002111 |
| GO:0032501 | multicellular organismal process | 802/1685 | 2.1589E-06 | 0.002132 |
| GO:0042221 | response to chemical | 513/1685 | 0.0001 | 0.018405 |
| GO:0042246 | tissue regeneration | 17/1685 | 0.0002 | 0.027754 |
| GO:0042692 | muscle cell differentiation | 55/1685 | 0.0003 | 0.037022 |
| GO:0043403 | skeletal muscle tissue regeneration | 11/1685 | 0.0003 | 0.044909 |
| GO:0043503 | skeletal muscle fiber adaptation | 4/1685 | 4.4531E-05 | 0.011269 |
| GO:0045165 | cell fate commitment | 46/1685 | 1.9621E-05 | 0.006707 |
| GO:0048018 | receptor ligand activity | 79/1656 | 2.4643E-06 | 0.002111 |
| GO:0051272 | positive regulation of cellular component movement | 84/1685 | 7.8914E-06 | 0.003597 |
| GO:0051493 | regulation of cytoskeleton organization | 73/1685 | 0.00012 | 0.020485 |
| GO:1902903 | regulation of supramolecular fiber organization | 51/1685 | 0.0004 | 0.047617 |
| GO:1904888 | cranial skeletal system development | 15/1685 | 0.0003 | 0.044566 |
| GO:2001046 | positive regulation of integrin-mediated signaling | 5/1685 | 6.6284E-05 | 0.014261 |
| **Cases vs. control smokers:** |  |  |  |  |
| GO:0003729 | mRNA binding | 23/81 | 1.0069E-05 | 0.001057 |
| GO:0010608 | posttranscriptional regulation of gene expression | 23/81 | 3.0028E-10 | 4.69E-08 |
| GO:0016441 | posttranscriptional gene silencing | 22/84 | 4.4483E-15 | 1.62E-12 |
| GO:0016442 | RISC complex | 23/81 | 7.5451E-17 | 6.64E-15 |
| GO:0016458 | gene silencing | 23/81 | 1.5066E-13 | 3.29E-11 |
| GO:0031047 | gene silencing by RNA | 22/84 | 1.2005E-14 | 3.28E-12 |
| GO:0031332 | RNAi effector complex | 23/81 | 7.5451E-17 | 6.64E-15 |
| GO:0035194 | posttranscriptional gene silencing by RNA | 23/81 | 4.3205E-15 | 1.62E-12 |
| GO:0035195 | gene silencing by miRNA | 23/81 | 3.2204E-15 | 1.62E-12 |
| GO:0040029 | regulation of gene expression, epigenetic | 23/81 | 7.5803E-13 | 1.38E-10 |
| GO:1903231 | mRNA binding involved in posttranscriptional gene silencing | 23/84 | 2.5252E-08 | 5.3E-06 |
| GO:1990904 | ribonucleoprotein complex | 23/84 | 1.9793E-08 | 1.16E-06 |

Gene ratio – number of significant genes identified in the data set as a ratio of the total number of genes in a pathway.

Twenty five of 37 uniquely enriched pathways in cases were in comparison to the non-smoking control group, of which 20 were in the GO domain of biological process (BP) and five in the domain molecular function (MF). For BP domain the significant terms were: GO:0001501, GO:0007186, GO:0007200, GO:0007399, GO:0008154, GO:0009888, GO:0009953, GO:0010454, GO:0032501, GO:0042221, GO:0042246, GO:0042692, GO:0043403, GO:0043503, GO:0045165, GO:0051272, GO:0051493, GO:1902903, GO:1904888, and GO:2001046. For MF domain the significant terms were: GO:0005125, GO:0005198, GO:0019958, GO:0030545, and GO:0048018. The remaining 12 of the 37 pathways uniquely enriched in cases were in comparison to cancer-free smokers. These were in BP (N=7), MF (N=2) and cellular component (CC; N=3) domains. For BP domain the significant terms were: GO:0010608, GO:0016441, GO:0016458, GO:0031047, GO:0035194, GO:0035195, and GO:0040029. For MF domain the significant terms were: GO:0003729 and GO:1903231. For CC domain the significant terms were: GO:0016442, GO:0031332, and GO:1990904.

## 4. Discussion

Various subtypes of circulating cfRNA have been tested in plasma for early-stage detection of NSCLC. Building upon these studies, we performed a comprehensive analysis of circulating plasma cfRNA using next generation sequencing technologies to expand the repertoire of non-invasively measurable NSCLC signatures. We identified 2357 cfRNA enriched in 123 pathways in those with a diagnosis of NSCLC compared to control groups consisting of cancer-free smokers and non-smokers. Nearly half of the detected cfRNA were transcripts of protein coding genes, and 251 of 2357 cfRNA (10.65%) conformed to previously reported differentially expressed genes in primary tumor biopsies from NSCLC patients. A majority (174 of 251) of these replicated transcripts were protein coding genes, while the rest were previously reported miRNA and other non-coding RNAs. In fact, two of the snoRNAs - SNORD115-41 and SNORD12, were previously reported in NSCLC tissue biopsies by our group [22].

Importantly, our pilot study used a workflow that can be easily adopted to develop a clinical assay for profiling cfRNA using plasma volumes smaller than that were reported elsewhere[56]. The archived plasma samples were derived from whole blood collected in standard 3-6 ml EDTA collection tubes routinely used in clinical care. Processing of small amounts of plasma (approximately 1.5ml) yielded less than 5ng of total cfRNA, and library preparation with enrichment and sequencing was carried out for efficient identification of cfRNA. Our methodology produced 200 to 350 million of sequence reads per sample, with over 80% of reads mapping to exonic regions of the reference, comparable to what is reported in methods that required much higher volumes of plasma [57].

Although identifying biomarker signatures associated with NSCLC was not the primary objective of this proof-of-concept pilot study that sought to test the potential of an NGS-based method for comprehensive detection of circulating cfRNA in plasma, we further evaluated cfRNA of the 251 genes to explore potential candidates for future NSCLC associated biomarker development studies. We first searched for cfRNA that were differentially expressed in plasma samples from NSCLC patients (regardless of the subtypes) compared to both smokers with benign PNs and non-smokers. Our results indicated 15 genes that included six protein coding, six miRNA and three other non-coding genes. Twelve of the 15 genes had low inter-patient variability (i.e., CV <50%) for cfRNA expression. These included five cf-mRNA (ARHGEF18, RAB38, PDE4DIP, BLID, and SRXN1), four cf-miRNA (MIR135A2, MIR193B, MIR617, MIR125B2), and all three of the other non-coding genes (SNORD115-41, SNORD12, SNHG1). Notably, cfRNA for the two snoRNAs SNORD115-41 and SNORD12 genes which we have reported previously[22], were not detectable in any NSCLC sample, but were present in both control groups with low inter-subject variability, confirming their potential role as plasma biomarkers of NSCLC. Furthermore, identifying protein coding genes (i.e., cf-mRNA) with low inter-patient variability was particularly significant as studies on circulating cf-mRNA are relatively sparse compared to miRNA or other non-coding genes. Thus, we tested the differential expression of the five cf-mRNA associated with NSCLC in a different cohort of NSCLC patients, smokers with benign PN, and non-smokers using quantitative RT-PCR. Our results indicated differential expression of cfRNA for ARHGEF18, PDE4DIP, and SRXN1 genes, but not RAB38 and BLID. The ARHGEF18 (Rho/Rac Guanine Nucleotide Exchange Factor 18) also known as P114-RhoGEF activates the downstream gene RhoA which is important for cell migration and tumor progression[58, 59]. Song et al. showed that ARHGEF18 gene was upregulated in squamous-cell carcinoma compared to adenocarcinoma or non-tumor tissue, and significantly associated with lung cancer lymph node metastasis[31]. In line with these findings, we detected an upregulation of ARHGEF18 in our discovery cohort (Figure 3 and Table 2), but a downregulation in the validation sample (Figure 4). It is possible that the reversal in direction of expression levels in the validation cohort occurred due to suboptimal qRT-PCR assay conditions as described below, rather than due to biological differences. The PDE4DIP (Phosphodiesterase 4D Interacting Protein) that anchors phosphodiesterase in centrosomes[35] was shown to co-express with the endogenous tumor suppressor gene THBS1, and high expression levels of PDE4DIP were associated with improved survival rates in adenocarcinoma patients[34]. Additionally, an exome-wide study of peripheral blood samples identified a frame-shift mutation in PDE4DIP of cancer patients but not in cancer-free family members suggesting a possible association of PDE4DIP with development of squamous cell lung cancer[35]. The SRXN1 (Sulfiredoxin 1) - another phosphodiesterase 4D anchoring protein, was found to be upregulated in lung cancer cell lines A549 and 95D and 75 NSCLC tissues compared to the adjacent non-tumor tissue. In our study both PDE4DIP and SRXN1 were downregulated in the discovery and validation cohorts[39]. More studies are needed to characterize the directionality associated with clinical characteristics of NSCLC development and progression.

Our pilot study has several limitations. First, biological factors such as gender and age have been shown to play a major role in the development and prognosis of lung cancers [60]. For example, women smokers have greater risk for developing lung cancer compared to men who smoke, presumably due to underlying genetic and other biological differences between men and women [61, 62]; the AC subtype predominates in women, whereas SCC are more common in men [63]; and individuals aged 65 and older are at greater risk of developing lung cancers[60]. The over-representation of samples from male patients as compared to the two control groups, and the modest sample size in this pilot project limited our ability to explore moderating effects of these biological factors on our findings. This is particularly true of the subtype analyses that revealed 452 differentially expressed cfRNA between AC and SCC groups, and 1,075 between stages I and II that consisted of small numbers of patients. Second, both groups of smokers - with and without cancer, were significantly older than the non-smoking control group in the discovery cohort. The larger numbers of DEGs that we detected in comparisons of NSCLC patients and non-cancer smokers with non-smokers may possibly have arisen from confounding effects of age-related alterations in expression of genes (see Figure 1.1). However, we were able to validate three out of five selected genes tested in an independent cohort with a balanced age distribution between comparison groups. Third, because of lack of information on stable endogenous reference gene(s) for the normalization of qRT-PCR data for circulating mRNA, we conducted validation analyses for the subset of five genes without the use of an endogenous control. Systematic analyses are urgently required to identify candidate genes with stable expression levels of cf-mRNA across samples for continued research on cf-mRNA analysis in NSCLC. Perhaps large RNA-seq data sets on circulating transcriptomes in plasma from NSCLC patients could facilitate such analyses. Fourth, we were not able to test the tissue specificity of the identified cfRNA because of the unavailability of lung tissue biopsies from the included participants for direct comparisons with plasma cfRNA. Nevertheless, we have utilized two control groups to adjust for confounding effects of smoking on cfRNA expression levels and applied conservative statistical thresholds of 5% FDR and a minimum of 2-fold change difference in expression level between conditions to reduce false positive findings. Furthermore, the fact that we were able to detect cfRNA of hundreds of previously reported RNA transcripts from primary NSCLC biopsies, is indeed promising.

In summary, we have presented transcriptome-wide cfRNA profiling using small volumes of plasma providing a framework for developing a non-invasive (blood-based) assay for potential early detection, diagnosis, and monitoring of NSCLC to facilitate high rates of patients able to receive curative surgical resections. Further studies are required for evaluation of our methodology and its clinical application.

## Data Availability

All data produced in the present study are available upon reasonable request to the authors

## Supplementary Materials

The following supporting information can be downloaded at: […],

Supplementary Table 1: Differentially expressed cfRNA

Supplementary Table 2: Genes included in enriched pathways

Supplementary Figure 1: qRT-PCR analysis of spike-in controls for cfRNA isolation across samples

## Author Contributions

Conceptualization, C.S., A.C.S., F.J., K.M. and S.F.; methodology, C.S., A.C.S., F.J., and S.F.; formal analysis, A.C.S. and C.M.; resources, X.X.; data curation, C.S., A.C.S., X.G., C.M. and J.C.; writing—original draft preparation, C.S.; writing—review and editing, A.C.S., F.J., K.M. and S.F.; supervision, C.S., A.C.S., F.J., and S.F.; funding acquisition, S.F.. All authors have read and agreed to the published version of the manuscript

## Institutional Review Board Statement

The study was conducted in accordance with the Declaration of Helsinki and approved by the Institutional Review Board (or Ethics Committee) of the University of Maryland Baltimore [UMB IRB protocol ID: HP-00040666] and the Veterans Affairs Maryland Health Care System [protocol ID: VA-00040666].

## Informed Consent Statement

Informed consent was obtained from all subjects involved in the study.

## Acknowledgments

Authors would like to thank Dan Gheba and Tara Kesteloot for their expert advice with developing the assays, John Sivinski for his assistance with sample processing, and Dr. Lisa Sadzewicz and Sandra Ott for advice and assistance with two-step sequencing.

## References

1. American Cancer Society. Facts & Figures 2022. American Cancer Society. Atlanta, Ga., 2022.

2. WHO global report on trends in prevalence of tobacco use 2000-2025. Fourth ed. 2021, Geneva: World Health Organization.

3. Li, Y., et al., Genome-wide interaction analysis identified low-frequency variants with sex disparity in lung cancer risk. Hum Mol Genet, 2022.

4. Besaratinia, A., A. Caceres, and S. Tommasi, DNA Hydroxymethylation in Smoking-Associated Cancers. Int J Mol Sci, 2022. 23(5).

5. Huang, Y., et al., Air Pollution, Genetic Factors, and the Risk of Lung Cancer: A Prospective Study in the UK Biobank. Am J Respir Crit Care Med, 2021. 204(7): p. 817–825.

6. Bade, B.C. and C.S. Dela Cruz, Lung Cancer 2020: Epidemiology, Etiology, and Prevention. Clin Chest Med, 2020. 41(1): p. 1–24.

7. Leduc, C., et al., Comorbidities in the management of patients with lung cancer. Eur Respir J, 2017. 49(3).

8. Campling, B.G., et al., Spontaneous smoking cessation before lung cancer diagnosis. J Thorac Oncol, 2011. 6(3): p. 517–24.

9. Siegel, R.L., et al., Cancer Statistics, 2021. CA Cancer J Clin, 2021. 71(1): p. 7–33.

10. Force., U.S.P.S.T., Final Update Summary: Lung Cancer: Screening.. https://www.uspreventiveservicestaskforce.org/Page/Document/UpdateSummaryFinal/lung-cancer-screening., July 2015.

11. Force, U.S.P.S.T., et al., Screening for Lung Cancer: US Preventive Services Task Force Recommendation Statement. JAMA, 2021. 325(10): p. 962–970.

12. Jonas, D.E., et al., Screening for Lung Cancer With Low-Dose Computed Tomography: Updated Evidence Report and Systematic Review for the US Preventive Services Task Force. JAMA, 2021. 325(10): p. 971–987.

13. Tanner, N.T., et al., Management of Pulmonary Nodules by Community Pulmonologists: A Multicenter Observational Study. Chest, 2015. 148(6): p. 1405–1414.

14. Pinzani, P., et al., Updates on liquid biopsy: current trends and future perspectives for clinical application in solid tumors. Clin Chem Lab Med, 2021. 59(7): p. 1181–1200.

15. Li, R.Y. and Z.Y. Liang, Circulating tumor DNA in lung cancer: real-time monitoring of disease evolution and treatment response. Chin Med J (Engl), 2020. 133(20): p. 2476–2485.

16. Gale, D., et al., Residual ctDNA after treatment predicts early relapse in patients with early-stage non-small cell lung cancer. Ann Oncol, 2022. 33(5): p. 500–510.

17. Larson, M.H., et al., A comprehensive characterization of the cell-free transcriptome reveals tissue- and subtype-specific biomarkers for cancer detection. Nat Commun, 2021. 12(1): p. 2357.

18. Sorber, L., et al., Circulating Cell-Free DNA and RNA Analysis as Liquid Biopsy: Optimal Centrifugation Protocol. Cancers (Basel), 2019. 11(4).

19. Muller, S., et al., Circulating MicroRNAs as Potential Biomarkers for Lung Cancer. Recent Results Cancer Res, 2020. 215: p. 299–318.

20. de Fraipont, F., et al., Circular RNAs and RNA Splice Variants as Biomarkers for Prognosis and Therapeutic Response in the Liquid Biopsies of Lung Cancer Patients. Front Genet, 2019. 10: p. 390.

21. Peng, W., et al., Diagnostic and Prognostic Potential of Circulating Long Non-Coding RNAs in Non Small Cell Lung Cancer. Cell Physiol Biochem, 2018. 49(2): p. 816–827.

22. Gao, L., et al., Genome-wide small nucleolar RNA expression analysis of lung cancer by next-generation deep sequencing. Int J Cancer, 2015. 136(6): p. E623–9.

23. https://www.illumina.com/products/by-type/sequencing-kits/library-prep-kits/rna-prep-enrichment.html.

24. Shetty, A.C., et al., CAVERN: Computational and visualization environment for RNA-seq analyses. 69th Annual Meeting. Houston, Texas: American Society of Human Genetics., 2019.

25. Kim, D., et al., Graph-based genome alignment and genotyping with HISAT2 and HISAT-genotype. Nat Biotechnol, 2019. 37(8): p. 907–915.

26. Anders, S., P.T. Pyl, and W. Huber, HTSeq--a Python framework to work with high-throughput sequencing data. Bioinformatics, 2015. 31(2): p. 166–9.

27. Benjamini, Y. and Y. Hochberg, Controlling the False Discovery Rate: A Practical and Powerful Approach to Multiple Testing. Journal of the Royal Statistical Society. Series B (Methodological), 1995. 57(1): p. 289–300.

28. Yamashita, S., et al., Combination of p53AIP1 and survivin expression is a powerful prognostic marker in non-small cell lung cancer. J Exp Clin Cancer Res, 2009. 28: p. 22.

29. Ye, T., et al., Chemokine CCL17 Affects Local Immune Infiltration Characteristics and Early Prognosis Value of Lung Adenocarcinoma. Front Cell Dev Biol, 2022. 10: p. 816927.

30. Yang, J., et al., Circular RNA CHST15 Sponges miR-155-5p and miR-194-5p to Promote the Immune Escape of Lung Cancer Cells Mediated by PD-L1. Front Oncol, 2021. 11: p. 595609.

31. Song, C., et al., Expression of p114RhoGEF predicts lymph node metastasis and poor survival of squamous-cell lung carcinoma patients. Tumour Biol, 2013. 34(3): p. 1925–33.

32. Hsieh, J.J., et al., RAB38 is a potential prognostic factor for tumor recurrence in non-small cell lung cancer. Oncol Lett, 2019. 18(3): p. 2598–2604.

33. Chang, J.W., et al., Comparison of genomic signatures of non-small cell lung cancer recurrence between two microarray platforms. Anticancer Res, 2012. 32(4): p. 1259–65.

34. Weng, T.Y., et al., Differential Expression Pattern of THBS1 and THBS2 in Lung Cancer: Clinical Outcome and a Systematic-Analysis of Microarray Databases. PLoS One, 2016. 11(8): p. e0161007.

35. Li, S., et al., Sequencing study on familial lung squamous cancer. Oncol Lett, 2015. 10(4): p. 2634–2638.

36. Wang, H., et al., Overexpression of ELF3 facilitates cell growth and metastasis through PI3K/Akt and ERK signaling pathways in non-small cell lung cancer. Int J Biochem Cell Biol, 2018. 94: p. 98–106.

37. Li, Z., et al., miRNA-124 modulates lung carcinoma cell migration and invasion. Int J Clin Pharmacol Ther, 2016. 54(8): p. 603–12.

38. Wei, Q., et al., Sulfiredoxin-Peroxiredoxin IV axis promotes human lung cancer progression through modulation of specific phosphokinase signaling. Proc Natl Acad Sci U S A, 2011. 108(17): p. 7004–9.

39. Zhou, J., et al., Identification of SRXN1 and KRT6A as Key Genes in Smoking-Related Non-Small-Cell Lung Cancer Through Bioinformatics and Functional Analyses. Front Oncol, 2021. 11: p. 810301.

40. Zhang, Y., et al., ARID1A is downregulated in non-small cell lung cancer and regulates cell proliferation and apoptosis. Tumour Biol, 2014. 35(6): p. 5701–7.

41. Wang, N. and T. Zhang, Downregulation of MicroRNA-135 Promotes Sensitivity of Non-Small Cell Lung Cancer to Gefitinib by Targeting TRIM16. Oncol Res, 2018. 26(7): p. 1005–1014.

42. Hu, F., et al., Lung adenocarcinoma resistance to therapy with EGFRtyrosine kinase inhibitors is related to increased expression of cancer stem cell markers SOX2, OCT4 and NANOG. Oncol Rep, 2020. 43(2): p. 727–735.

43. Choi, K.H., et al., Dual-strand tumor suppressor miR-193b-3p and -5p inhibit malignant phenotypes of lung cancer by suppressing their common targets. Biosci Rep, 2019. 39(7).

44. She, K., et al., miR-193b availability is antagonized by LncRNA-SNHG7 for FAIM2-induced tumour progression in non-small cell lung cancer. Cell Prolif, 2018. 51(1).

45. Chen, W.J., et al., MicroRNA-153 expression and prognosis in non-small cell lung cancer. Int J Clin Exp Pathol, 2015. 8(7): p. 8671–5.

46. Yuan, Y., et al., Suppression of AKT expression by miR-153 produced anti-tumor activity in lung cancer. Int J Cancer, 2015. 136(6): p. 1333–40.

47. Zhang, W., et al., Expressions of connexin 32 and 26 and their correlation to prognosis of non-small cell lung cancer. Ai Zheng, 2009. 28(2): p. 173–6.

48. Shan, N., et al., MiR-153 inhibits migration and invasion of human non-small-cell lung cancer by targeting ADAM19. Biochem Biophys Res Commun, 2015. 456(1): p. 385–91.

49. Kim, H.K., et al., miR-592 and miR-552 can distinguish between primary lung adenocarcinoma and colorectal cancer metastases in the lung. Anticancer Res, 2014. 34(5): p. 2297–302.

50. Huang, S.P., et al., Downregulation of miR-125b-5p and Its Prospective Molecular Mechanism in Lung Squamous Cell Carcinoma. Cancer Biother Radiopharm, 2022. 37(2): p. 125–140.

51. Wang, J., et al., Expression and clinical evidence of miR-494 and PTEN in non-small cell lung cancer. Tumour Biol, 2015. 36(9): p. 6965–72.

52. Wang, M., et al., High expression of kinesin light chain-2, a novel target of miR-125b, is associated with poor clinical outcome of elderly non-small-cell lung cancer patients. Br J Cancer, 2015. 112(5): p. 874–82.

53. Zhou, Q., et al., A novel lncRNA-miRNA-mRNA competing endogenous RNA regulatory network in lung adenocarcinoma and kidney renal papillary cell carcinoma. Thorac Cancer, 2021. 12(19): p. 2526–2536.

54. Tan, J., et al., Integrative Analysis of Three Novel Competing Endogenous RNA Biomarkers with a Prognostic Value in Lung Adenocarcinoma. Biomed Res Int, 2020. 2020: p. 2837906.

55. Shi, S.L. and Z.H. Zhang, Long non-coding RNA SNHG1 contributes to cisplatin resistance in non-small cell lung cancer by regulating miR-140-5p/Wnt/beta-catenin pathway. Neoplasma, 2019. 66(5): p. 756–765.

56. Mullins, K., et al., Proof of Concept: detection of cell free RNA from EDTA plasma in patients with lung cancer and non-cancer patients. medRxiv, 2022: p. 2022.08.12.22278721.

57. Rasmussen, M., et al., RNA profiles reveal signatures of future health and disease in pregnancy. Nature, 2022. 601(7893): p. 422–427.

58. Terry, S.J., et al., Stimulation of cortical myosin phosphorylation by p114RhoGEF drives cell migration and tumor cell invasion. PLoS One, 2012. 7(11): p. e50188.

59. Kim, M., et al., p114RhoGEF governs cell motility and lumen formation during tubulogenesis through a ROCK-myosin-II pathway. J Cell Sci, 2015. 128(23): p. 4317–27.

60. Leiro-Fernandez, V., et al., Predicting delays in lung cancer diagnosis and staging. Thorac Cancer, 2019. 10(2): p. 296–303.

61. Hellyer, J.A. and M.I. Patel, Sex disparities in lung cancer incidence: validation of a long-observed trend. Transl Lung Cancer Res, 2019. 8(4): p. 543–545.

62. Molina, A.J., et al., Trends in Lung Cancer Incidence in a Healthcare Area. Arch Bronconeumol, 2015. 51(11): p. e53–5.

63. Pesch, B., et al., Cigarette smoking and lung cancer--relative risk estimates for the major histological types from a pooled analysis of case-control studies. Int J Cancer, 2012. 131(5): p. 1210–9.

